# Force of infection: A determinant of vaccine efficacy?

**DOI:** 10.1101/2021.01.21.21250235

**Authors:** David C. Kaslow

## Abstract

Vaccine efficacy (VE) can vary in different settings. Of the many proposed setting-dependent determinants of VE, force of infection (FoI) stands out as one of the most direct, proximate, and actionable. As highlighted by the COVID-19 pandemic, modifying FoI through non-pharmaceutical interventions (NPIs) use can significantly contribute to controlling transmission and reducing disease incidence and severity absent highly effective pharmaceutical interventions, such as vaccines. Given that NPIs reduce the FoI, the question arises as to if and to what degree FoI, and by extension NPIs, can modify VE, and more practically, as vaccines become available for a pathogen, whether and which NPIs should continue to be used in conjunction with vaccines to optimize controlling transmission and reducing disease incidence and severity.

## Introduction

Lower apparent vaccine efficacy (VE) in low resource settings, when compared to VE observed in high resource settings, has been reported for several pathogens, most notably poliovirus, typhoid, and rotavirus.^1–5^ Observed VE also varied when evaluating a malaria vaccine candidate in different parasite transmission settings.^6–8^ Numerous economic, social, and biological factors have been proposed to explain these setting-dependent variations in VE.^3,9–11^ Many, if not most, of the proposed economic and social determinants of VE, such as, country income status, living conditions, access to healthcare, appear to act indirectly and non-specifically on VE; whereas many but not all biological factors, such as co-infections, malnutrition, and enteropathy, presumably act directly and proximally on VE. More practically, identification of direct and proximal determinants of setting-dependent VE that hold the promise of actionable intervention(s) seem a most urgent need in efforts to enhance and/or sustain VE.

The COVID-19 pandemic has highlighted the contribution of non-pharmaceutical interventions (NPIs) in controlling transmission and reducing disease incidence and severity,^12^ particularly in the absence of highly effective pharmaceutical interventions, such as vaccines. NPIs also contribute to controlling other major human diseases, including use of condoms for HIV/AIDS^13^, bed nets for malaria^14^, and hand washing for diarrhea^15^. By reducing the number of (susceptible) individuals effectively contacted by each (infected) person, e.g., through physical barriers, distancing, and masking, NPIs reduce λ, the force of infection (FoI) (see **Box 1**, Glossary of Key Terms). As vaccines become available for a pathogen, the question arises as to if and which NPIs should continue to be used, if not prioritized.^16^ This then begs the broader use-inspired scientific question, as raised previously:^8^ after optimizing the vaccine immunogen, formulation, dose level, and regimen, what remaining determinants of vaccine efficacy (VE) are amenable to intervention? More specifically, given the role of NPIs in reducing the FoI, if and to what degree is FoI, and by extension NPIs, a determinant of VE?

### Interrogating the potential relationship of force of infection and setting-dependent vaccine efficacy

A two-step approach was taken to interrogate the potential relationship between FoI and VE. The first explored three mathematical scenarios of VE as a function of various FoI settings. The second followed up on the decades old observations of lower apparent oral poliovirus vaccine (OPV)^1^ and typhoid vaccine^5^ VE in low resource settings when compared to high resource settings by assessing the correlation between the incidence of disease in the placebo population (as a surrogate of FoI in the study population) and the observed VE in different geographical settings. Recent Phase 3 studies of malaria and rotavirus vaccine candidates across a number of settings, including low and high resource settings,^6,17^ provided data for assessing if and how FoI might be a determinant of VE.

Both the thought experiment of setting-dependent VE of a hypothetical vaccine and the retrospective analyses of rotavirus and malaria Phase 3 efficacy results make a multitude of assumptions that limit the robustness and soundness of any conclusions. For simplicity, factors previously shown or hypothesized to influence transmission, susceptibility, VE, and/or FoI, such as, country income status, age, underlying medical conditions, co-infections, access to healthcare, seasonality, NPI use, spreading events, and strain differences across different settings, were excluded from consideration in both the hypothetical VE or observed VE analyses.

Given these significant limitations in the analyses, the primary goal of the present study was not to provide a definitive answer to the questions of if and to what degree FoI determines VE in different settings. Rather the goal of these analyses was to continue to raise the awareness of the potential impact of FoI on VE,^8,18^ and to prompt prospective studies designed to assess if and how NPIs might reduce FoI and enhance VE when vaccines are introduced and scaled up. Ultimately well-designed studies that directly evaluate the potential relationship of FoI and setting-dependent VE will provide the evidence needed for well-informed policy recommendations on the continued use or not of NPIs during vaccine introduction and scale-up.

### Three scenarios of the potential mathematical consequences of force of infection on setting-dependent vaccine efficacy

The potential effects of FoI on the level of VE were explored in three mathematical scenarios: 1) VE_constant_, where VE is independent of FoI; 2) VE_linear_, where VE decreases linearly as a function of increasing FoI; and, 3) VE_natural log_, where VE decreases logarithmically as FoI. As noted above, multiple simplifying assumptions were made when considering the mathematical consequences of FoI on VE, including homogeneity in the population with respect to a number of factors, including pathogen transmission, host susceptibility to infection and disease, FoI over time in a specific setting, and protective immunity as a result of vaccination across settings.

With these simplifying assumptions in mind, equations that define the three mathematical scenarios (see **Box 2**, Vaccine efficacy as a function of Force of infection) are shown graphically in **Fig. 1**, using the example of a hypothetical vaccine that has a maximum VE of 83% studied under conditions of FoI that vary across three orders of magnitude, from 0.03 to 3.50 infections/person-year. While other mathematical relationships between VE and FoI may be considered, these three equations seemed a reasonable starting point from which to interrogate observed data from Phase 3 VE studies conducted in multiple epidemiological settings.

**Fig 1.**
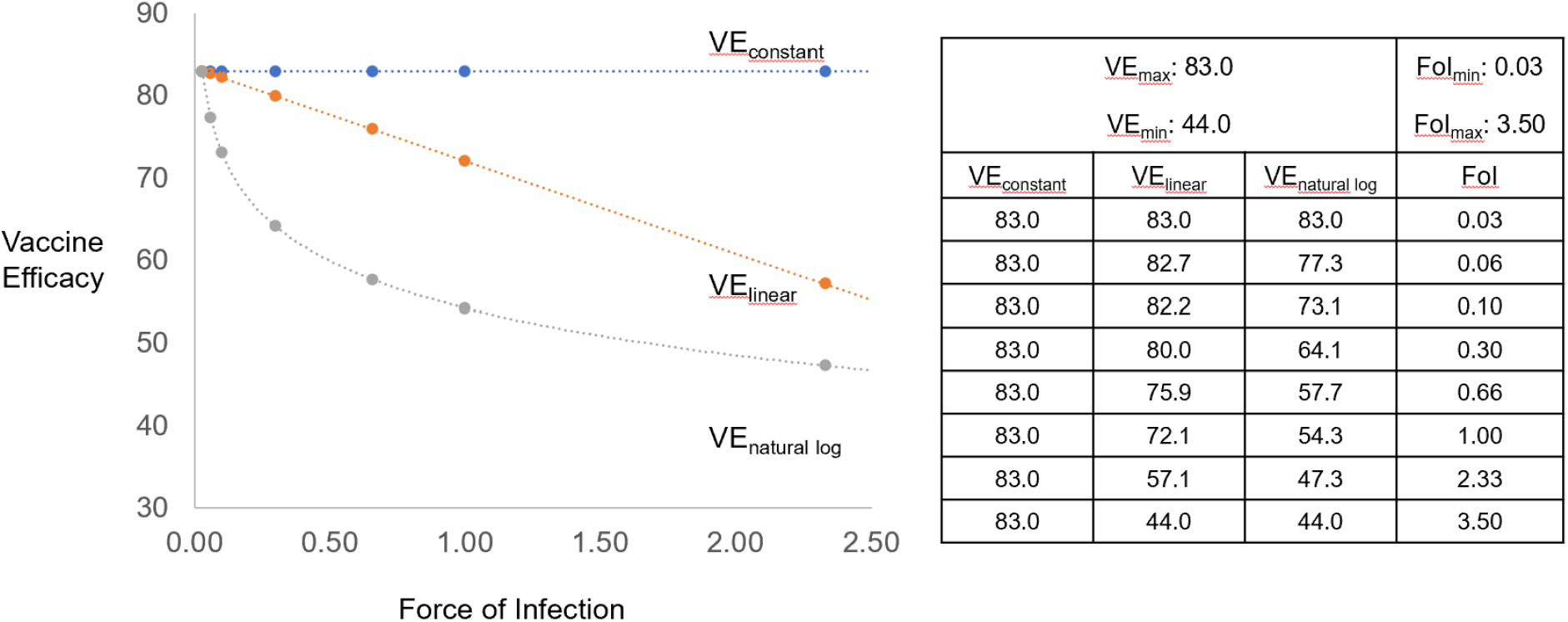
Vaccine Efficacy (VE) as a function of Force of Infection (FoI) for hypothetical vaccine. Equations that define three mathematical scenarios (see **Box 2**, Vaccine efficacy as a function of force of infection) are shown graphically, using as an example a hypothetical vaccine with a maximum vaccine efficacy (VE_max_) of 83% and minimum VE (VE_min_) of 44.0% studied under conditions of force of infection (FoI) that vary across three orders of magnitude, from a minimum FoI (FoI_min_) 0.03 to a maximum FoI (FoI_max_) of 3.50 infections/person-year.

### Empiric evidence of force of infection on observed setting-dependent vaccine efficacy

Results from recent placebo-controlled Phase 3 studies of vaccine candidates for two diverse pathogens, *Plasmodium falciparum* and rotavirus, provided a database to determine which, if any, of the three mathematical scenarios best explained any setting-dependent differences in VE. In addition to the assumptions mentioned above, several additional assumptions noted below facilitated the analyses of these multi-setting VE studies.

First and foremost, the analyses of both pathogens assumed that the intent-to-treat (ITT) incidence of the most sensitive definition of the mildest disease end point in the youngest age cohort in the placebo arm best served as an internal Phase 3 study surrogate of λ, the force of infection. The validity of this assumption relies upon several other assumptions, including the absence of any significant herd effect (see **Box 1**, Glossary of Key Terms) on the control from the vaccinated arm of the Phase 3 study. The rationale for making this herd effect assumption, typically also assumed for the control used in estimating VE in the context of Phase 3 efficacy studies, relies upon: 1) the relatively small proportion of the total population in the study setting enrolled in the vaccinated group in the Phase 3 study; and, 2) the timing of incident disease in the control group relative to eliciting herd immunity and reaching the herd immunity threshold (see **Box 1**, Glossary of Key Terms) in the study population.

A third key assumption relied upon a comparison of trendlines from the three mathematical scenarios described above to the closest fit trendline of observed VE (VE_observed_) as a function of observed setting FoI (FoI_observed_, incidence in the placebo group) to determine if and how VE varied as a function of FoI. In this regard, because the Phase 3 VE results for both pathogens were known a priori to vary by epidemiologic setting, the posterior probability was low of selecting the VE_constant_ mathematical scenario to categorize VE_observed_ as a function of FoI_observed_. As noted below for each specific analysis, the observed trendline may not necessarily reflect a statistically significant association between VE_observed_ and FoI_observed_, as assessed by a regression analysis.

### Malaria parasite vaccine efficacy and force of infection

A single pivotal Phase 3 VE study (NCT00866619) enrolled 15,459 participants in two age categories (young children aged 5–17 months and infants aged 6–12 weeks at the time of enrollment) across 11 clinical research sites in seven African countries (one site in Burkina Faso, Gabon, Malawi, and Mozambique; two sites in Ghana and Tanzania; and three sites in Kenya). The trial assessed, as a primary aim, VE of a three-dose regimen of RTS,S/AS01_E_ against clinical malaria over 12 months follow-up.^7^ In the per-protocol population of the 5–17 months age category, VE_observed_ was 51.3% (95% CI: 47.5 - 54.9; p-value <.0001) with a VE_observed_ range from 83.0% (95% CI: 37.2 - 95.4; p-value 0.0079) in a low parasite transmission site (Kilifi, Kenya) to 44.0% (95% CI: 36.8 - 50.3; p-value <.0001) in a high parasite transmission site (Nanoro, Burkina Faso) (see Annex 6 table 23, Ref 19). As noted above, the intent-to-treat (ITT) incidence of the more sensitive secondary definition of clinical malaria in the control group of infants aged 6–12 weeks at the time of enrollment (see Annex 7 table 177, Ref 20) served as the internal Phase 3 study FoI_observed_, the surrogate of λ in the analyses.

The best fit trendline analysis of VE_observed_ as a function of FoI_observed_ revealed a logarithmic relationship (**Fig. 2**, Observed VE) with an R^2^ of 0.807. Regression analysis of VE_observed_ as a function of ln FoI_observed_ revealed a Significance F of 0.006. Using the VE_natural log_ equation (**Box 2**), the observed VE_max_, VE_min_, FoI_max_, FOI_min_ and the FoI_observed_ from each site generated a logarithmic relationship between the calculated site-specific VE and FoI_observed_ (**Fig 2**, Calculated VE). These analyses suggest that FoI functions as a determinant of RTS,S/AS01_E_ VE.

**Fig 2.**
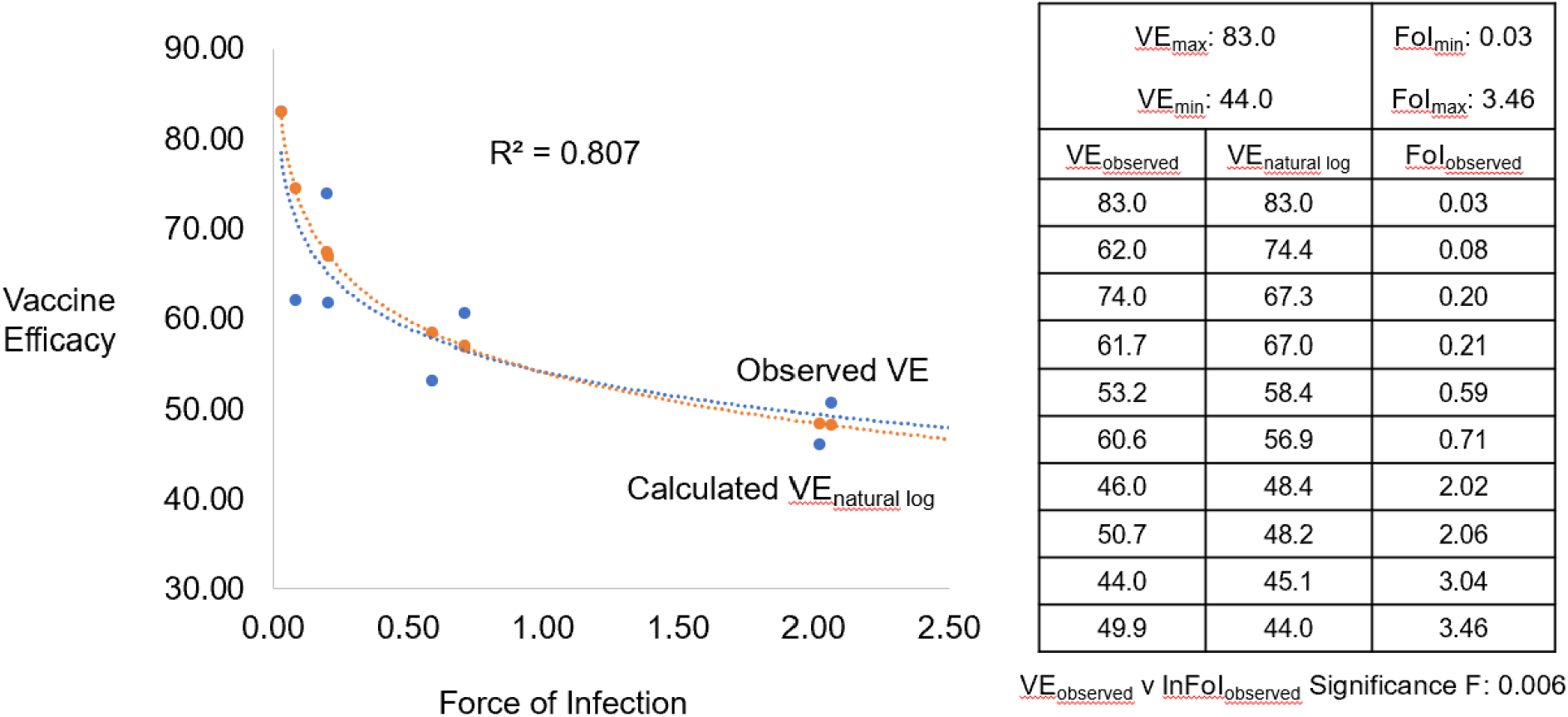
Vaccine Efficacy (VE) as a function of Force of Infection (FoI) for malaria vaccine. Best fit trendline analysis of observed vaccine efficacy (VE_observed_) as a function of observed force of infection (FoI_observed_) is shown as a logarithmic relationship (blue dotted line) with a R^2^ of 0.807. Significance F of 0.006 from a regression analysis of VE_observed_ as a function of ln FoI_observed_ in the embedded table is shown. Using the VE_natural log_ equation (see **Box 2**, Vaccine efficacy as a function of force of infection), the observed VE_max_, VE_min_, FoI_max_, FOI_min_ and FoI_observed_ were used to calculate the VE_natural log_ in the embedded table and the calculated VE_natural log_ shown graphically (orange dotted line).

### Rotavirus vaccine efficacy and force of infection

Multiple Phase 3 studies of two rotavirus vaccines, RV1 (Rotarix^®^) and RV5 (RotaTeq^®^), evaluated VE in diverse epidemiologic settings.^17^ In comparison to the analyses conducted for malaria VE, the analyses of VE_observed_ as a function of FoI_observed_ for rotavirus vaccines was complicated by the evaluation of two different vaccine candidates, with two different regimens, in several different clinical protocols. Some of the Phase 3 studies conducted in low resource settings did not collect data on the incidence of rotavirus gastroenteritis (RVGE) of any severity. The analyses excluded these studies due to the absence of an intent-to-treat incidence of any severity RVGE in the placebo group to serve as a surrogate of λ. The analyses also excluded data from countries in which the placebo group had no or just a single case of RVGE of any severity. From those studies that collected sufficient incidence of any severity RVGE in the placebo group, an Analysis of Variance failed to detect a statistically significant difference (p-value = 0.749) when categorizing FoI_observed_ by 2020 World Bank country income classifications (i.e., upper-v upper middle-v lower middle/lower-income country)^21^ (**Table 1**).

**Table 1.**
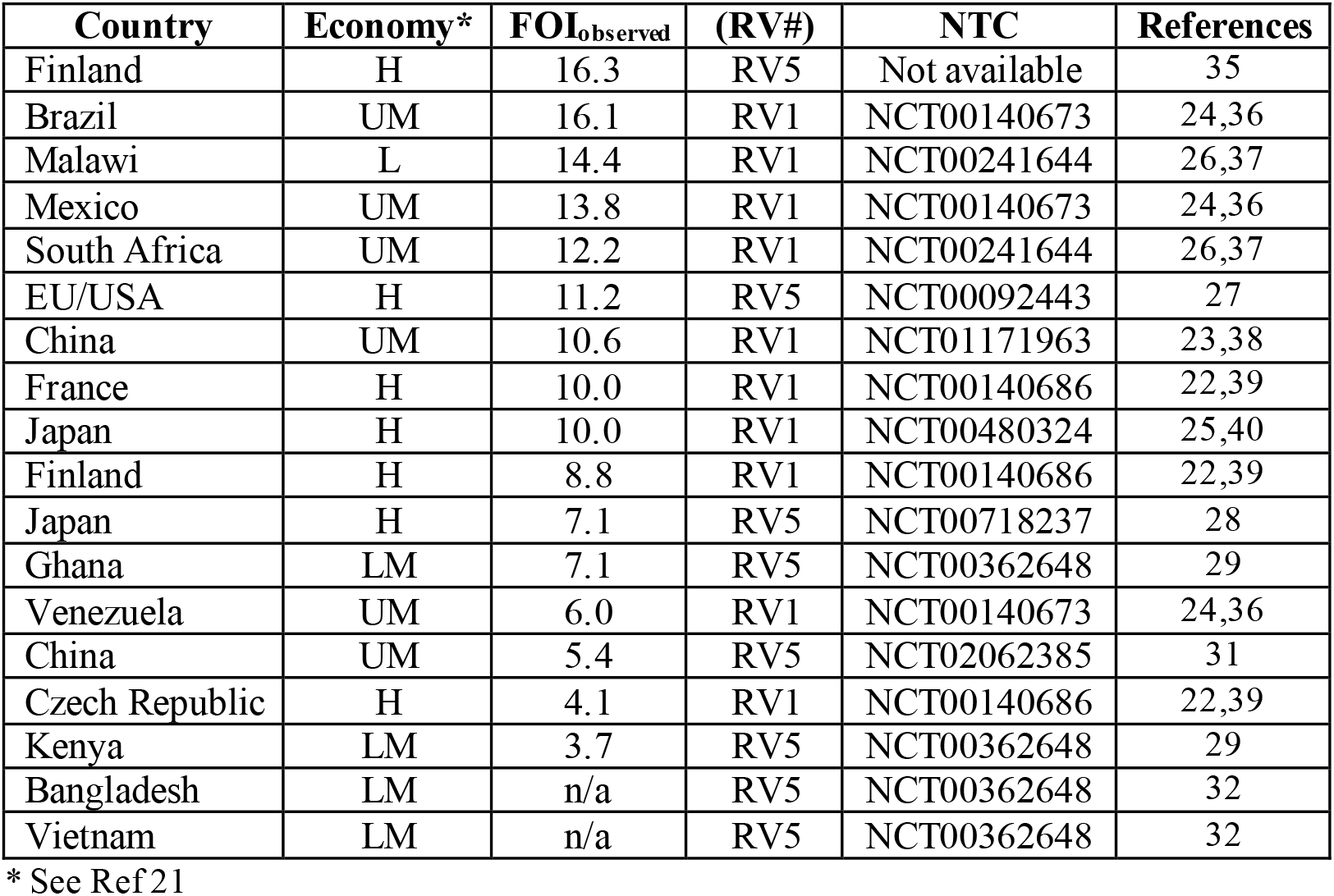

| **Country** | **Economy\*** | **FOI<sub>observed</sub>** | **(RV#)** | **NTC** | **References** |
| --- | --- | --- | --- | --- | --- |
| Finland | H | 16.3 | RV5 | Not available | 35 |
| Brazil | UM | 16.1 | RV1 | NCT00140673 | 24,36 |
| Malawi | L | 14.4 | RV1 | NCT00241644 | 26,37 |
| Mexico | UM | 13.8 | RV1 | NCT00140673 | 24,36 |
| South Africa | UM | 12.2 | RV1 | NCT00241644 | 26,37 |
| EU/USA | H | 11.2 | RV5 | NCT00092443 | 27 |
| China | UM | 10.6 | RV1 | NCT01171963 | 23,38 |
| France | H | 10.0 | RV1 | NCT00140686 | 22,39 |
| Japan | H | 10.0 | RV1 | NCT00480324 | 25,40 |
| Finland | H | 8.8 | RV1 | NCT00140686 | 22,39 |
| Japan | H | 7.1 | RV5 | NCT00718237 | 28 |
| Ghana | LM | 7.1 | RV5 | NCT00362648 | 29 |
| Venezuela | UM | 6.0 | RV1 | NCT00140673 | 24,36 |
| China | UM | 5.4 | RV5 | NCT02062385 | 31 |
| Czech Republic | H | 4.1 | RV1 | NCT00140686 | 22,39 |
| Kenya | LM | 3.7 | RV5 | NCT00362648 | 29 |
| Bangladesh | LM | n/a | RV5 | NCT00362648 | 32 |
| Vietnam | LM | n/a | RV5 | NCT00362648 | 32 |

\* See Ref 21

For RV1, results from 10 countries in five independent Phase 3 studies^17,22–26^ (see **Table 1**) met the above FoI_observed_ criteria for interrogation. The best fit trendline analysis of VE_observed_ as a function of FoI_observed_ revealed a linear relationship (**Fig. 3a** upper line, Observed VE) with an R^2^ of 0.3892 and regression analysis with a Significance F of 0.158. The VE_observed_ of 94.9% in one setting (Mexico) with FoI_observed_ of 13.79 appeared to be a significant outlier. Reanalysis absent the data from Mexico revealed a linear relationship (**Fig. 3a** middle line, Observed VE), with an R^2^ of 0.6264 and regression analysis Significance F of 0.0449. Using the VE_linear_ equation (**Box 2**), the observed VE_max_, VE_min_, FoI_max_, FOI_min_ and the FoI_observed_ from each of the 10 countries generated a linear relationship between the calculated site-specific VE and FoI_observed_ (**Fig. 3a** lower line, Calculated VE). These analyses suggest that FoI may function as a determinant of RV1 VE.

**Fig 3.**
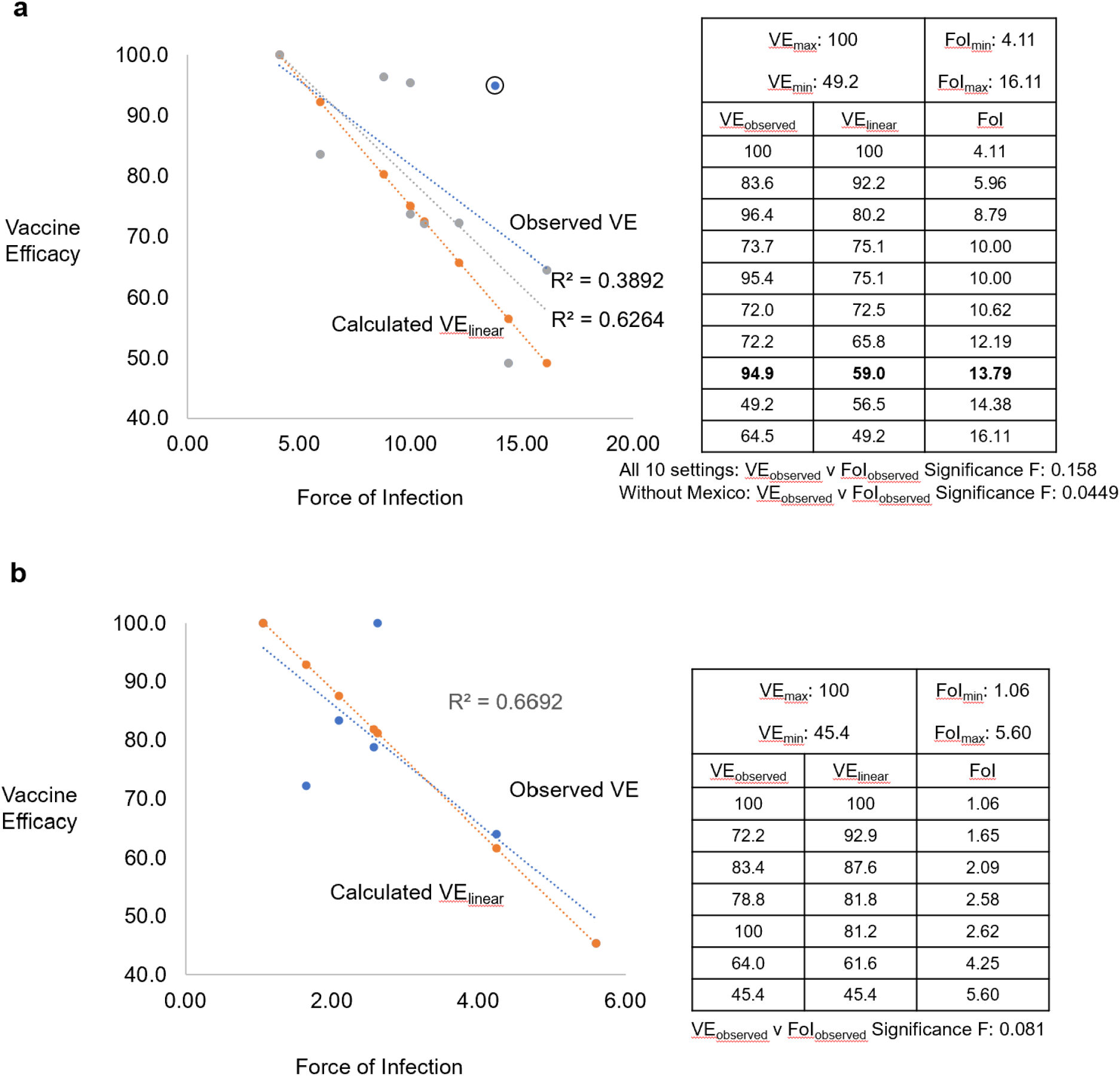
Vaccine Efficacy (VE) as a function of Force of Infection (FoI) for rotavirus vaccines. **a** RV1: Best fit trendline analysis of observed vaccine efficacy (VE_observed_) as a function of observed force of infection (FoI_observed_) is shown as a linear relationship for all 10 countries (blue dotted line) and for 9 countries (exclusion of the outlier, encircled blue dot; gray dotted line) with a R^2^ of 0.3892 and 0.6264, respectively. Significance Fs of 0.158 and 0.0449 from a regression analysis of VE_observed_ as a function of FoI_observed_ in the embedded table are shown. Using the VE_natural log_ equation (see **Box 2**, Vaccine efficacy as a function of force of infection), the observed VE_max_, VE_min_, FoI_max_, FOI_min_ and FoI_observed_ were used to calculate the VE_natural log_ in the embedded table and the calculated VE_natural log_ shown graphically (orange dotted line). **b** RV5: Best fit trendline analysis of observed vaccine efficacy (VE_observed_) as a function of observed force of infection (FoI_observed_) is shown as a linear relationship (blue dotted line) with a R^2^ of 0.6692. Significance F of 0.081 from a regression analysis of VE_observed_ as a function of FoI_observed_ in the embedded table is shown. Using the VE_natural log_ equation (see **Box 2**, Vaccine efficacy as a function of force of infection), the observed VE_max_, VE_min_, FoI_max_, FOI_min_ and FoI_observed_ were used to calculate the VE_natural log_ in the embedded table and the calculated VE_natural log_ shown graphically (orange dotted line).

For RV5, results from 5 settings in three independent Phase 3 studies^17,27–31^ (see **Table 1**) met the above FoI_observed_ criteria for interrogation. The best fit trendline analysis of VE_observed_ as a function of FoI_observed_ revealed an independent relationship (data not shown) with an R^2^ of -0.215 and regression analysis Significance F of 0.9838. Interrogating results from 7 settings in five independent Phase 3 studies^17,27–32^ (see **Table 1**) by using the incidence of SRVGE in the placebo group as the FoI_observed_ and surrogate of λ in the analyses, the best fit trendline analysis of VE_observed_ as a function of FoI_observed_ revealed a linear relationship (**Fig 3B**, Observed VE) with an R^2^ of 0.6692 and regression analysis Significance F of 0.081. Using the VE_linear_ equation (**Box 2**), the observed VE_max_, VE_min_, FoI_max_, FOI_min_ and the FoI_observed_ from each of the 7 settings in the reanalysis generated a linear relationship between the calculated site-specific VE and FoI_observed_ (**Fig 3B** lower line, Calculated VE). These analyses suggest that FoI may function as a determinant of RV5 VE, when the FoI_observed_ in the analyses is the incidence of SRVGE in the placebo group.

## Conclusion

Of the many proposed determinants of setting-dependent VE, FoI provides one of the most direct, mechanistically proximate potential determinants. For many but not all pathogens, modifying the FoI provides one of the most actionable interventions to enhance or sustain VE. While improving indirect or distal VE determinants, such as poverty, gut pathology, co-infections, and malnutrition, could significantly enhance efforts to control and eliminate simultaneously many pathogens, implementing interventions that effectively mitigate these VE determinants is complex and not immediately achievable. In contrast, modifying the FoI through the concomitant use of affordable, accessible, available, acceptable, and sustainable NPIs provides a proximate and actionable approach to optimizing VE. Considering and then prospectively verifying the speculation that introduction or continued optimal use of NPIs in an effort to reduce the FoI and thereby enhance or sustain VE, respectively, upon vaccine rollout seems prudent and, in the context of a pandemic, quite urgent.

## Data Availability

All data in the data in the manuscript are included in the manuscript.

## Text boxes

### Box 1.

#### Glossary of key terms

**Force of infection**

Rate at which susceptible individuals in a population acquire an infectious disease in that population, per unit time. It is also known as the incidence rate or hazard rate.^33^

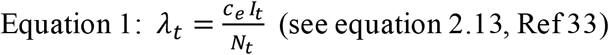

Where λ_t_ = force of infection at time t, c_e_ = number of individuals effectively contacted by each person per unit time, I_t_ = number of infected in the population at time t, N_t_ = number in the population at time t

**Efficacy**

The direct protection provided by vaccination against a defined clinical endpoint; it excludes any indirect (herd) effect.^33^

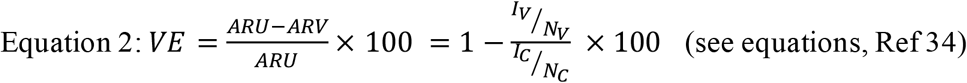

Where VE = vaccine efficacy, ARU = attack rate in the unvaccinated population, ARV = attack rate in the vaccinated population, I_V_ = number of infected in the vaccinated population, N_V_ = number in the vaccinated population, I_U_ = number of infected in the unvaccinated population, and N_U_ = number in the unvaccinated population

**Herd immunity**

The proportion of a population immune to infection or disease.^2,33^

**Herd immunity threshold**

The proportion of the population required to be immune in the population for the infection incidence to reach steady state, i.e., the infection level is neither growing nor declining. To eliminate an infection in the population, the proportion of the population that is immune to infection must exceed this threshold value.^33^

**Herd effect**

The reduction in the rate of infection or disease in the unimmunized portion of a population as a result of immunizing a proportion of the population.^2^ It is also known as indirect effect.

### Box 2.

#### Vaccine efficacy as a function of Force of infection

The following equations define mathematical relationships between Vaccine Efficacy (VE) and Force of Infection (FOI) shown in Figure 1, when the relationship of VE is: 1) independent of FOI (VE_constant_); 2) linear to FOI (VE_linear_); or 3) logarithmic to FOI (VE_natural log_):

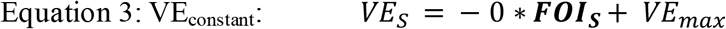

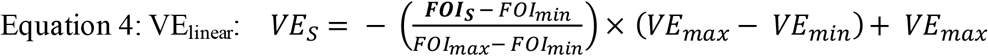

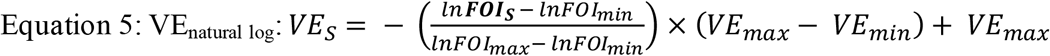

Where: *VE*_*S*_ is the VE in setting *S, VE*_*max*_ is the highest observed VE, and *VE*_*min*_ is the lowest observed VE.

And, where *FOI*_*S*_ is the FOI in setting *S, FOI*_*min*_ is lowest observed FOI, and *FOI*_*max*_ is the highest observed FOI.

